# Are current NEWS2 clinical response thresholds optimised for a general in-patient population?

**DOI:** 10.1101/2020.09.12.20136804

**Authors:** Tanya Pankhurst, Elizabeth Sapey, Helen Gyves, Felicity Evison, Suzy Gallier, Georgios Gkoutos, Simon Ball

## Abstract

**Background:** The National Early Warning Score 2 (NEWS2) is mandated in acute hospital trusts in England. Assessment of the implications of this policy across an unselected in-patient population has been limited.

**Objective:** Evaluate NEWS2 performance in an acute, in-patient, population by relating potential costs and benefits of specific alerting thresholds 24 hours prior to a composite outcome event (unplanned intensive care admission or death).

**Methods:** All in-patient spells between Nov 2018 - Jul 2019 in a single acute hospital in the UK were analysed. Standardised Early Warning Score(SEWS) and NEWS2 data acquisition was from the electronic health record (EHR). Existing SEWS alert thresholds were maintained. The performance of NEWS2 and SEWS threshold score against the composite outcome was assessed. A single clinical review cost (€129.50) was used to model the whole system cost of triggered responses at different NEWS2 thresholds.

**Results:** In patients ≥24 hours post-admission, a mean daily rate of progression to the outcome event was 1.95/1000. An increase in alert threshold from NEWS2 ≥5 to ≥6 reduced the proportion that would trigger clinical review from 10.0% to 5.3% per day. This was associated with the false negative rate at threshold increasing from 1.13/1000 patients to 1.36/1000. A simple resource model allowing one triggered clinical response every 24 hours defined an incremental cost per patient benefiting =€26,463, equating to 18 additional healthcare professionals per 1000 patients to deliver clinical response to an additional 0.23 patients/day benefitting.

**Conclusion:** The low event rate across the whole in-patient population, moderate performance of a single NEWS2 score and associated resource requirements mean that in any resource limited setting, ‘rules-based’ unmodified NEWS2 response thresholds may divert clinical resource and focus.

**Summary Box:** *What is already known on this topic?:* NEWS2 is mandated as an early warning score for all NHS acute hospitals in England. There are associated graded clinical response recommendations progressing to urgent clinical review at a NEWS2 score ≥5. Evaluation of the clinical and health economic implications of applying these recommendations across a whole in-patient population has been limited.

*What this study adds:* This is the first study to report NEWS2 alert thresholds in a longitudinal hospital cohort and to model the cost of implementing a key threshold NEWS2 ≥5 for all in-patients. For in-patients ≥24 hours post-admission, approximately 10% trigger at least one NEWS2 score ≥5 per day (excluding those on intensive care or following a palliative care decision). The corresponding daily adverse event rate was 0.19% (admission to intensive care or death). The real world performance of current NEWS2 thresholds, using the observed classification performance, event rate and a response cost derived from the literature, identifies high incremental costs of increasing NEWS2 sensitivity.

## Introduction

The use of early warning scores (EWs) to harmonise and integrate physiological parameters into a single actionable output on presentation to hospital has been widely advocated. In 2017, the Royal College of Physicians (RCP) published an updated National Early Warning Score (NEWS), referred to as NEWS2 (1). NEWS2 is a scoring system based upon six physiological parameters. It is associated with specific clinical response recommendations with a step change occurring at a threshold NEWS2 score ≥5, which requires an urgent clinical response (1). In 2019, National Health Service England (NHSE) mandated the implementation of NEWS2 in acute hospital trusts and widened its application to include screening for sepsis (2). As digital maturity within the NHS grows, electronic health records (EHRs) are increasingly able to calculate EWs and automatically mandate an urgent clinical response in real time throughout an individual’s in-patient stay, widening the use of NEWS2 beyond the acute presentation in which it was developed (3-9).

The RCP’s urgent clinical response recommendations define an overhead arising from increased frequency of observation and the need for urgent clinical review. Although there are few estimations of the monetary costs, one study has suggested this is €129.50 (approximately £112) per patient (10). The cost of responding to patients with NEWS2 ≥5 is not just economic but includes diversion of clinical resource away from other patients with NEWS2 <5. Given limited direct evidence that EWS’s improve hard outcomes (11, 12), any new indication requires careful evaluation, to identify risks as well as benefits that may emerge from changes in organisational and professional behaviours. The real-world effectiveness of NEWS2 is consequently determined, not simply by its classification performance (described by its Receiver Operator Characteristic (ROC)) but the prevalence of meaningful outcomes and costs of responding to threshold scores.

We hypothesised that alerts based upon current thresholds would cause a high number of mandated clinical reviews across the whole acute hospital in-patient population in, despite a relatively low rate of adverse outcome events. This might represent a significant resource and opportunity cost. In this study we report a single centre, real-world evaluation of the relationship between NEWS2 score and outcome in an unselected acute hospital population, currently screened with SEWS via a rules-based automated electronic alert system (13). The performance of NEWS2 was assessed across the entire in-patient setting, where evidence to inform actionable thresholds and specify clinical responses is limited. A cost model was then applied to the outcome prediction of different NEWS2 thresholds.

## Methods

The Queen Elizabeth Hospital Birmingham (QEHB) is an adult acute hospital in England with 1269 beds including 100 level 2/3 intensive care (ICU) beds. The EHR at QEHB (PICS, Birmingham Systems) contains time-stamped, structured records that includes demography, location, time of admission and discharge, physiological measurements supporting NEWS2 and SEWS scores and non-resuscitation orders (a “do not attempt cardiopulmonary resuscitation” (DNACPR) order and palliative care pathway) as well as investigational requests and results and interventions.

As part of a change assessment programme (UHB clinical audit: CARMS-15850), data collection underpinning NEWS2 was initiated, whilst electronic alerting continued to use established SEWS thresholds and response recommendations (13). Admissions were identified as emergency or elective from the hospital provider spell administrative data return admission method code (Table 1). All in-patient spells between 00.00 on 01/11/2018 and 23.59 on 31/7/2019 were analysed. An adapted Consort diagram is shown in Supplementary figure S1a. Those in whom NEWS2 was recorded were organised into datasets permitting cross-sectional and longitudinal analysis.

**Table 1.** Demographic and Clinical characteristics of patients. The demographics, mode of admission and admitting speciality on first admission, of the 52,214 patients that were subject to NEWS2 analysis, contributing 36,182 elective and 42,249 emergency admissions

|  | Elective | Emergency |
| --- | --- | --- |
| Number of patients with one or more included admission | 22,538 | 29,122 |
| Mean age / years (at time of first admission) | 53.9 ± 18.1 | 56.2 ± 21.7 |
| Number of male patients (percentage) | 12,142<br>(53.9%) | 13,800<br>(47.4%) |
| ***Admitting speciality on first admission*** |  |  |
| General Medicine | 1545<br>(6.9%) | 20,174<br>(69.3%) |
| General Surgery | 2150 (9.5%) | 3061 (10.5%) |
| Trauma & Orthopaedics | 2307 (10.2%) | 1486 (5.1%) |
| Neurosurgery | 1866 (8.3%) | 639 (2.2%) |
| Urology | 1411 (6.3%) | 587 (2.0%) |
| Cardiology | 1821 (8.1%) | 469 (1.6%) |
| Clinical & Medical Oncology | 1263 (5.6%) | 549 (1.9%) |
| Ear, Nose and Throat (ENT) | 1401 (6.2%) | 389 (1.3%) |
| Plastic Surgery | 2138 (9.5%) | 304 (1.0%) |
| Maxillo-Facial Surgery | 849 (3.8%) | 252 (0.9%) |
| All others | 5787 (25.7%) | 1212 (4.2%) |
| Patients ineligible for analysis throughout their admission | 60 | 494 |
| Mean number of admissions / patient | 1.6 | 1.5 |

### Cross sectional analysis

All patients in a non-ICU bed at midday were identified on consecutive days. Each ‘patient-day’ from 12.00 to 11.59 was then analysed independently. Patients with a current palliative care pathway and DNACPR order (now referred to DNACPR only) as at midday and those on intensive care units (ICU) were excluded from that day’s cross-sectional EWS analysis. The population at midday was chosen as more representative of activity than the NHS administrative definition of a day in hospital, based upon bed occupancy at midnight.

### Longitudinal analysis

The first 14 days post-admission were assessed, for all patient spells across the study timeframe. Each spell was divided into consecutive 24-hour periods post-admission and in the primary analysis an Index NEWS2 score was defined as the first recorded in each period. An Index SEWS score was calculated alongside each Index NEWS2. EWS’s were excluded from analysis if at the time recorded the patient was in ICU, had a DNACPR in place, or for scores within 12 hours of admission if a DNACPR was instigated within those first 12 hours. If patients returned from ICU or the DNACPR was revoked, subsequent scores were then included in the analysis.

Supplementary Figure S1b summarises definitions used in longitudinal analysis, establishing a ‘patient-day’ variable that was treated independently. This design was used to avoid the association of a single outcome event with multiple NEWS2 scores, a potential source of bias if NEWS2 performance and the frequency of measurement are inter-dependent. Although this design minimises the effects of counting multiple NEWS2 scores, there remain a small number of events associated with two Index NEWS2 scores (10%) or with no Index NEWS2 score (3.5%). A sensitivity analysis, excluding double counted events in different ways had no meaningful effect on results.

### Outcomes

A composite outcome event was defined as an unplanned admission to HDU/ICU (type 1 and 2 of the critical care minimum dataset) or death of the patient in the 24 hours after the EWS score. This outcome definition is clinically meaningful and operationally relevant, in particular because 24 hours is the NHS minimum recommended frequency for senior medical review of in-patients (14).

### Elective versus unplanned admissions

For both cross-sectional and longitudinal cohorts, separate analyses were undertaken for Day 1 (first 24 hours following admission) and day 2-14 post admission, stratified by planned and unplanned admission as it was hypothesised that in the first 24 hours of elective admission, EWS’s would behave substantially differently than in other populations.

### EWS performance and cost model

The classification performance of EWS’s for the outcome event was evaluated by its ROC and associated c-statistic. ROC is a dimensionless measure of test performance suited to the comparison of EWS performance, but it can only partly inform the optimum NEWS2 threshold for escalation. The activity was therefore calculated across EWS to support real world evaluation of NEWS2 key trigger threshold implementation, as well as the corresponding false negative rate and Youden J statistic.

The RCP recommends: ‘hourly observation, urgent assessment by a clinician or team with core competencies in the care of acutely ill patients and the provision of clinical care in an environment with monitoring facilities’ (1), at a key threshold NEWS2 score ≥ 5. A cost of ‘medical emergency team’ assessment, reported from Holland in 2009 (€129.50 ~ £112) was used to model the cost of different key threshold scores (10), allowing for just one urgent clinical response/patient/day. The outcome event rate associated with a single NEWS2 score in longitudinal analysis was then used to calculate the incremental cost per outcome event in which a response would have been triggered at different NEWS2 thresholds.

The staffing resource required to meet RCP recommendations for NEWS2 ≥ 5 were calculated using the same assumptions as described (10). Here the time to respond to an alert call, staff grades of the response team and time taken for training requirements, rota maintenance twenty-four days a day, 7 days a week were quantified and costed. This provides an indicative cost for modelling pruposes and is not a full economic costing.

All statistical analyses were undertaken in STATA SE 15.1. Normally distributed variables are represented as mean ± standard deviation others as median and interquartile range. There were no adjustments for multiple comparisons and all p values are reported.

## Results

### NEWS2 in cross section analysis

The clinical and demographic details of the cross-sectional in-patient population are summarised in Table 1. 285,681 in-patient days were analysed. Of this population 79% had been in hospital ≥24 hours (Supplementary Table S2 in the online supplement). A NEWS2 score was recorded a mean of 3.7 times over the following 24 hours. This included 104 patients (median; IQR: 86-115) in whom no NEWS2 was recorded after midday, of whom 72 (median; IQR: 57-83) were discharged within the next 6 hours.

In any 24-hour period, a mean of 6.9 ± 1.1% of all NEWS2 scores were ≥ 5 and a mean of 9.8 ± 1.3% of patients generated at least one NEWS2 score ≥ 5, in whom the mean number of NEWS2 scores ≥ 5/patient/day = 2.6. These findings were not altered by excluding patients within 24 hours of admission (when a mean of 10.0 ± 1.4% of patients generated at least one NEWS2 score ≥ 5). If RCP recommended NEWS2 thresholds are used, of those already in a hospital bed at midday, a mean of 103 patients (9.8% of 1046) would require an urgent clinical response in every 24 hours period.

### Longitudinal analysis

The longitudinal analysis included 100,362 in-patient spells (see Supplementary Figure S1a). Of these, 21,931 did not include a NEWS2 score, in 21,046 of whom the length of stay was < 6 hours. Of the 78,431 spells with a NEWS2 score, 42,249 were emergency and 36,182 elective admissions.

The composite adverse event rate (death or unplanned ICU admission) was higher in the first 24 hours of emergency patient admission than the subsequent in-patient spell. (Daily adverse outcome rate for emergency admissions within 24 hours = 0.4%, after 24 hours = 0.2%, p<0.0001). The adverse event rate was lower in elective admissions (within 24 hours = 0.3%, after 24 hours =0.2%, p=0.008).

25,961 (71.8%) of the elective admissions had a length of stay <24 hours. Across all spells, an Index NEWS2 score was recorded 233,961 times in the first 14 days post-admission, during which time 1,051,599 NEWS2 scores were recorded in total.

In the longitudinal analysis, of all NEWS2 scores recorded within 14 days post-admission, 80,709 (7.7%) were ≥ 5, whilst of Index NEWS2 scores recorded in the first 14 days, 11,051 (4.7%) were ≥ 5.

The mean number of NEWS2 scores recorded/patient/day was higher for emergency admissions on day 1 (mean = 5.9 ± 0.1) than for all other days and routes of admission (mean = 4.2 ± 0.1). Figure 1 shows the probability that an Index NEWS2 score met different key trigger thresholds over the first 14 days post-admission. Day 1 emergency admissions had a high rate of Index NEWS2 score above these thresholds and elective admissions a low rate, which then converged. These differences confirmed our decision to analyse the ROC of the Index NEWS2 on day 1 separately from days 2-14 and stratified by emergency and elective admission. Index NEWS2 scores were associated with 615 composite outcome events, 307 from day 1.

**Figure 1.**
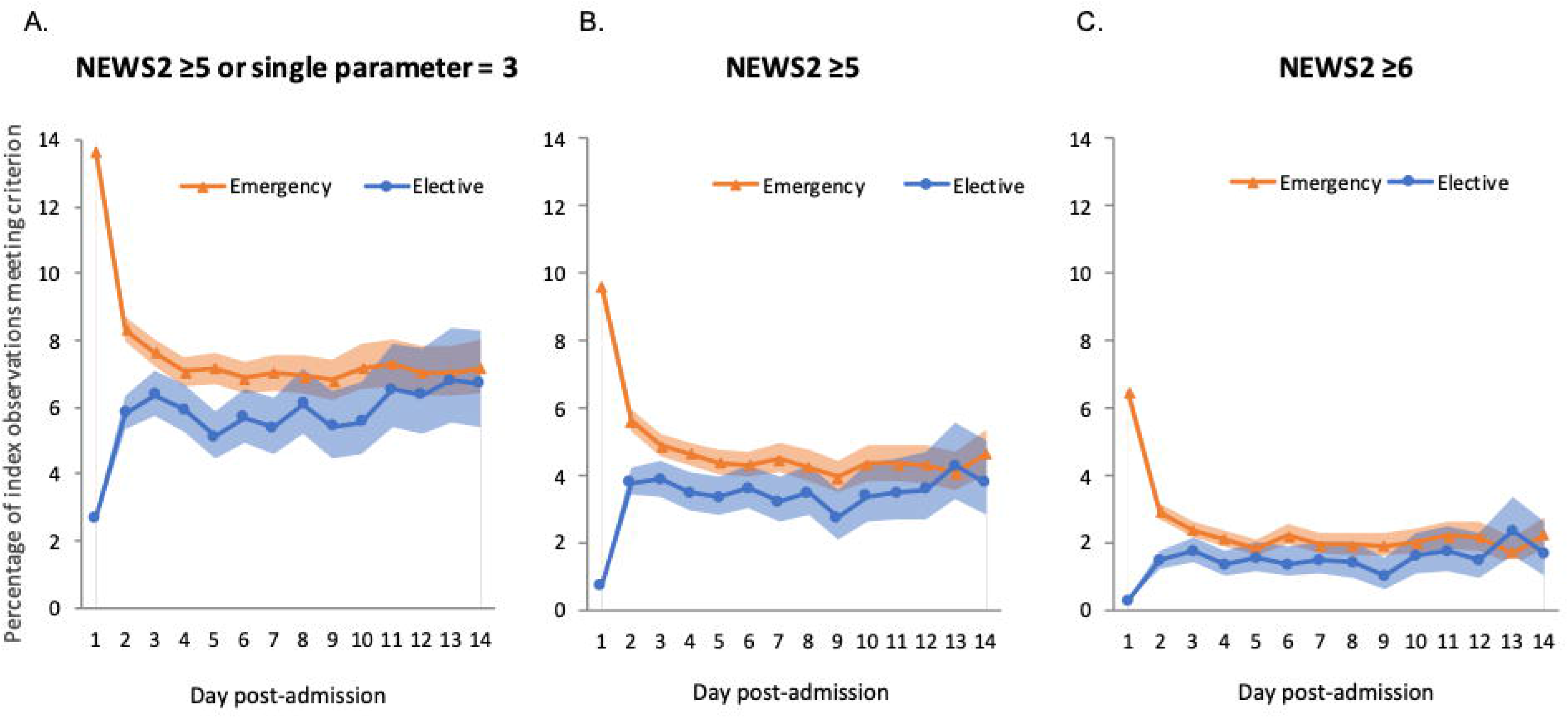
Changes in the percentage of patients meeting threshold scores. The percentage (daily mean with 95% confidence intervals) of patients in whom the Index NEWS2 met the heading’s threshold scores, is shown for emergency (▲) and elective admissions (●) on days 1 to 14 post-admission. (A) For all NEWS2 ≥ 5 or a single parameter =3. (B) For NEWS2 ≥ 5 and (C) For NEWS2 ≥ 6.

### Classification performance of NEWS2

The classification performance of Index NEWS2 and corresponding Index SEWS was calculated against the composite outcome event (Figure 2). The ROC of NEWS2 was superior to that of SEWS in all analyses. The superior performance of NEWS2 in emergency admissions extended out to 14 days post-admission (c statistic = 0.81). The Index NEWS2 performance on Day 1 of elective admission was not predictive for adverse outcomes (c statistic = 0.51). It almost always preceded a planned procedure occurring within 12 hours (94.7%) and was therefore recorded prior to any resulting physiological insult. A post-hoc analysis of the first NEWS2 recorded from 12 hours post elective admission gave a c-statistic = 0.63, whilst for day 2-14 post-admission the c-statistic = 0.70. Since elective admissions constitute 24.1% of the combined day 2-14 post-admission population, the performance of NEWS2 was good in the overall combined population (c-statistic = 0.77). The combined Day 2-14 population was used in subsequent analysis.

**Figure 2.**
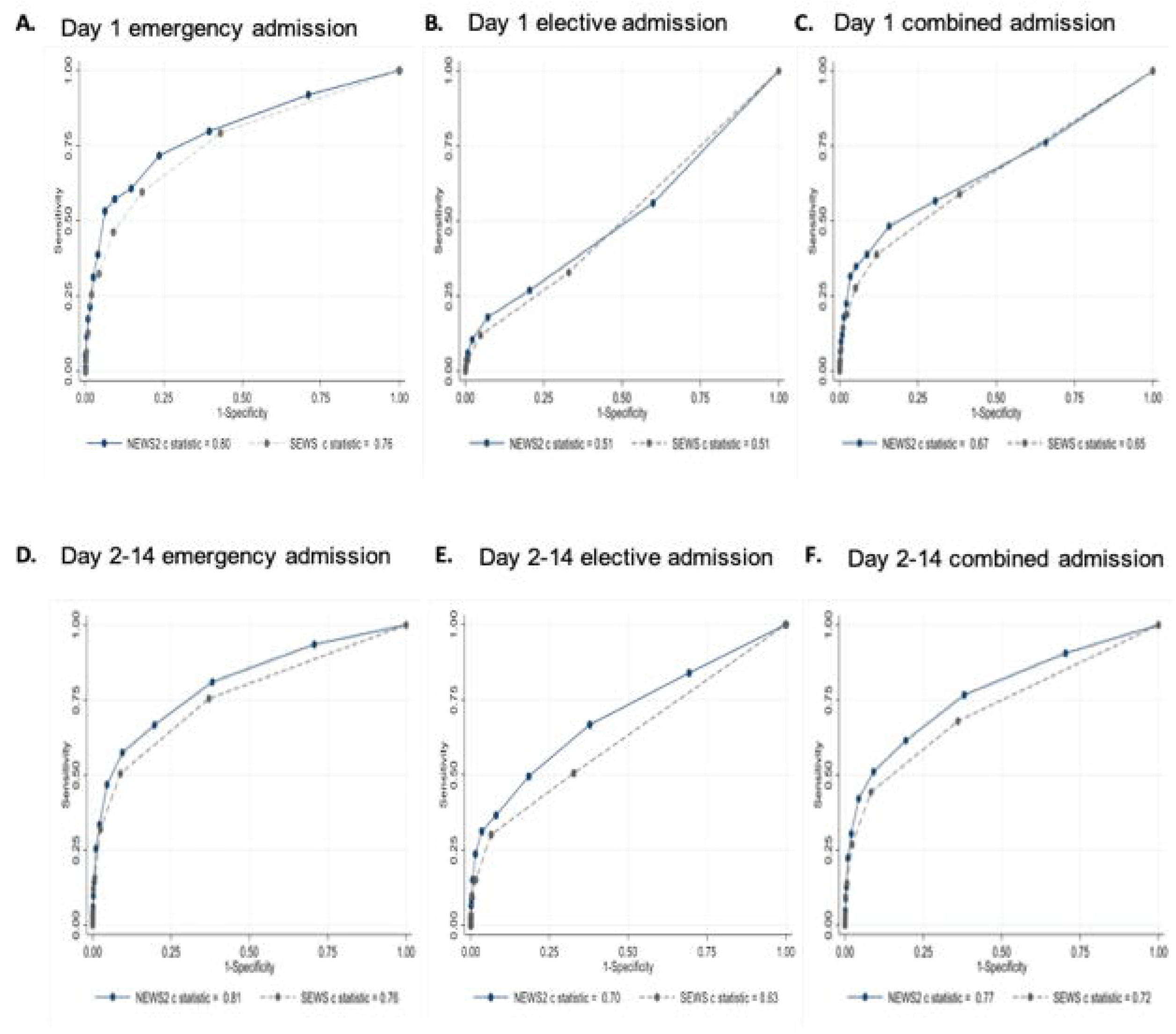
Receiver operating characteristic (ROC) for Index NEWS2 and SEWS. The ROC for index NEWS2 and SEWS for (A) Day 1 emergency admissions, (B) Day 1 elective admissions and (C) Day 1 combined (emergency + elective) admissions, (D) Day 2 – 14 emergency admissions, (E) Day 2-14 elective admissions and (F) Day 2-14 combined emergency + elective) admissions, discriminating the composite outcome event.

### Real world consequences of NEWS2 trigger thresholds

The activity across a range of Index NEWS2 and SEWS scores, along with the False Negative Rate and Youden J statistic, is shown for Day 1 of emergency admission (Table 2a) and the Day 2-14 combined admissions (Table 2b). The underlying data across all NEWS2 scores are summarised in supplementary Figure S3 and S4.

**Table 2a.**
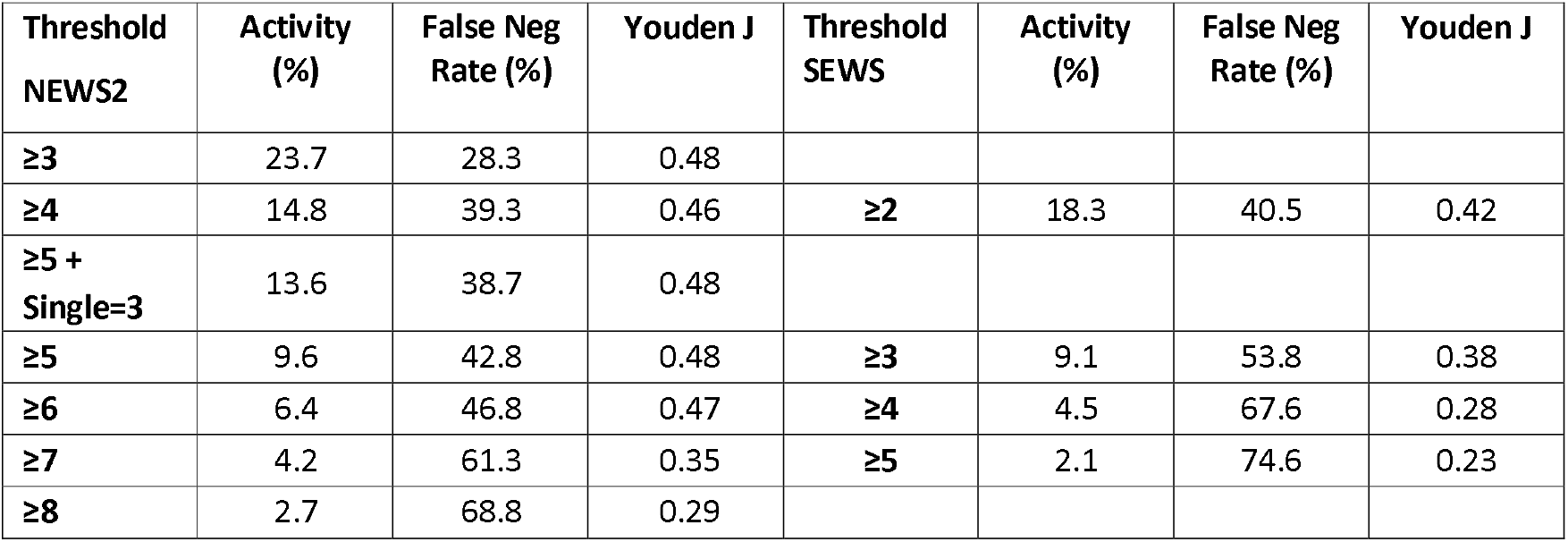
Performance of Index NEWS 2 and SEWS for Day 1 emergency admission: activity, false negative rate and Youden J statistic.

**Table 2b.**
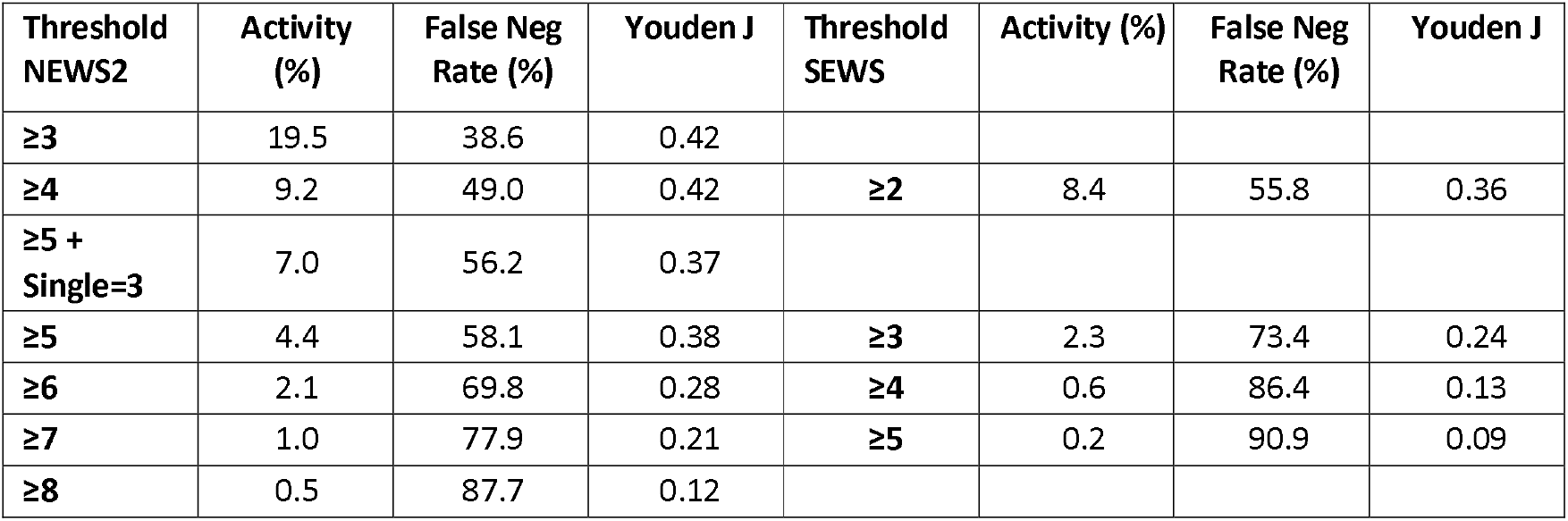
Performance of Index NEWS 2 and SEWS for Day 2-14 post admission: activity, false negative rate and Youden J statistic. a. data for the day one of emergency admission b. data for days 2 to 14 of all in-patient admissions The Index NEWS2 (or SEWS) score is the first recorded in each successive 24-hour period following admission. The composite outcome event is the first of death and unplanned admission to ICU, occurring in the 24-hour period following the Index NEWS2. Activity (column 2) is the percentage of Index NEWS2 or SEWS score meeting the threshold score = (TP+FP)/(TP+FP+TN+FN). (TP is true-positive, FP is false-positive, TN is truenegative FN is false-negative, where positive (P) is an EWS meeting the threshold criteria in column 1, and true (T) is occurrence of the composite outcome e vent). The false negative rate is the percentage of outcome events not meeting the threshold EWS = FN/(TP+FN). These data correspond to the activity plot in supplementary Figure 3 and raw data in supplementary Figure 4.

These data allow the real-world consequences of different NEWS2 key trigger threshold scores to be estimated. For the combined Day 2-14 population, Table 3 shows the estimated daily costs of responding to one NEWS2-triggered medical alert per patient per day, using an urgent clinical response cost of €129.50 with Table 3a describing the cost of implementation from day 2-14 and Table 3b the cost of implementation for Day 1 post emergency admission. The incremental cost per outcome event in which a response would have triggered at different threshold scores was then derived (10). For example, the cost of triggering at NEWS2 ≥ 5 vs NEWS2 ≥ 6 would be €26,463 (£22,886) per ensuing additional composite outcome event in which a response had been triggered. The generalisation of Index NEWS2 to any NEWS2 score, in the combined Day 2-14 population, is supported by analysis of a random selection of single daily NEWS 2 scores (supplementary Figure S5). A similar calculation was made for incident patients on day 1 post emergency admission (as shown in Table 3b) and a similar step increase in incremental cost below a threshold NEWS2 ≥6 was observed. Although the absolute number of additional events at Index NEWS2 = 4 and 5 were relatively low on day 1 (supplementary Fig S4), a sensitivity analysis suggests that this finding is robust.

**Table 3a.**
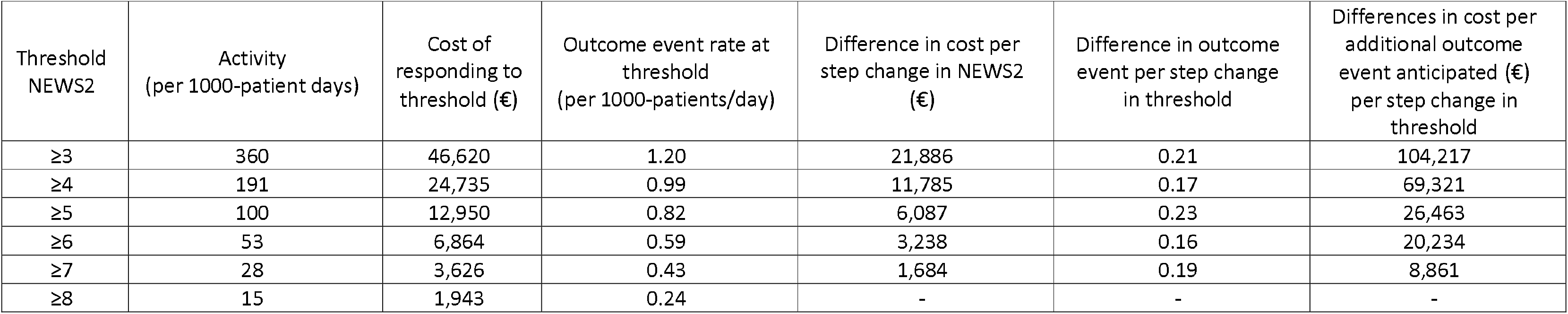
Performance of Index NEWS 2 – cost of implementation for the population Day 2-14 post admission. The activity and cost of responding to different key NEWS2 threshold scores (column 1), in the prevalent population at midday, who had been in hospital ≥24 hours, is shown per 1000 patients/day (columns 2-3). This assumes the cost of responding to threshold = €129.50. It allows for only one response per day (12.00 - 11.59), per patient triggering at the specified threshold NEWS2 score, irrespective of the number of NEWS2 scores triggered that day. The outcome event rate associated with a single NEWS2 score, at different thresholds, per 1000 patients/day is shown (column 4). Th e effects of single step reductions in threshold NEWS2 score on the cost per 1000 patients/day (column 5) and on the outcome event rate (column 6) at th reshold are shown. The effects of single step reductions in threshold NEWS2 score, on the nominal cost of responding per additional outcome event predicted is shown (column 7)

**Table 3b.**
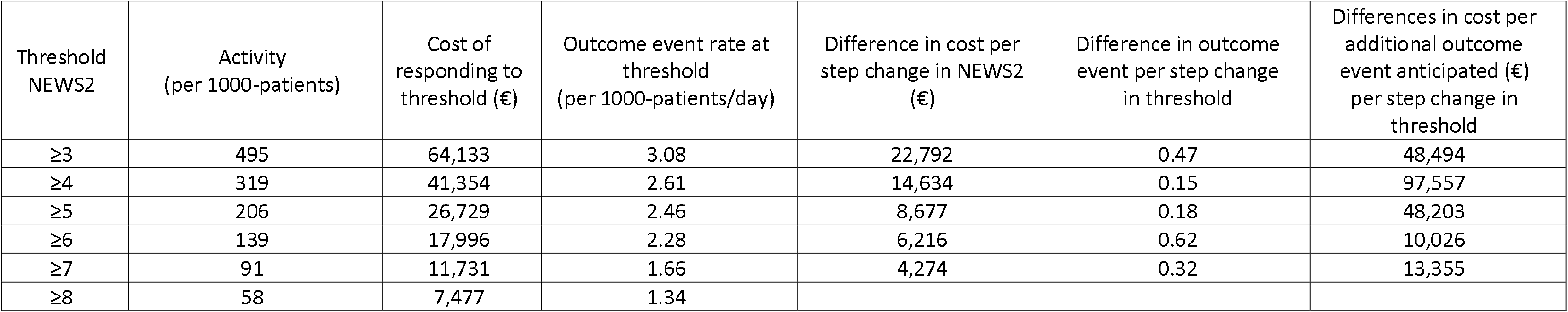
Performance of Index NEWS 2 – cost of implementation for Day 1 post emergency admission. The activity and cost of responding to different key NEWS2 threshold scores (column 1) in the first 24 hours following emergenc y admission, is shown per 1000 patients (columns 2-3). This assumes the cost of responding to threshold = €129.50(10). It allows for only one response in the day at the specified threshold NEWS2 score, irrespective of the number of NEWS2 scores triggered that day. The outcome event rate associated with a single NEWS2 score, at different thresholds, per 1000 patients is shown (column 4). The effects of single step reductions in threshold NEWS2 score on the cost per 1000 patients (column 5) and on the outcome event rate at threshold (column 6) are shown. The effects of single step reductions in threshold NEWS2 score, on the nominal cost of responding per additional outcome event predicted is shown (column 7)

Using the same report’s assumptions on staffing requirements to deliver a clinical response (10), based on a 40 hour working week and NHS standard terms and conditions for annual leave and study leave, an additional 18 healthcare professionals per 1000 patients, would be needed to provide reviews for all inpatients triggering at NEWS2 ≥5 compared to NEWS2 ≥ 6. This corresponds to an additional 0.23 patients/day being reviewed who then went on to sustain a composite outcome event. It is based on a simplified model in which only one alert triggers a clinical response in any 24 hour period.

## Discussion

This study provides a real-world evaluation of the implications of NEWS2 implementation across an in-patient spell. It describes the clinical and operational consequences of specific NEWS2 response thresholds and recommendations, arguably a more important consideration in determining the disposition of resource than marginal differences in EWS classification performance.

Standardised clinical pathways provide safe, efficient and equitable healthcare that can be easily understood, measured and improved (15). NEWS was established in a paper system to create an NHS wide ‘track and trigger’ system to realise such benefits during acute presentation and early events in hospital (1, 3). NEWS2 has not been so comprehensively evaluated and not all analyses report benefits, for example from the recognition of patients at risk of hypercapnic respiratory failure (16,17). There is a need for ongoing real world evaluation of NEWS2 (16-18), particularly following NHSE’s requirement that it be used to monitor all adults in acute hospitals (2).

In our real-world evaluation, the ROC of NEWS2 was superior to that of the currently used SEWS in all patient populations, for a standard, clinically meaningful, composite outcome event. Importantly, NEWS2 performance remained acceptable out to 14 days post admission. An initial conclusion from these results is that at QEHB, NEWS2 is superior to SEWS for cross-organisational monitoring of the whole in-patient population, not just acute admissions. However, the finding that NEWS2 is superior to SEWS does not identify the optimal threshold for clinical response and this is an important question when extending NEWS2 screening from acute admission units to the whole in-patient population, in which higher activity is accompanied by a lower event rate.

Our data show that a key threshold score does not emerge simply from the characteristics of the test. For example, the Youden J statistic in Day 1 emergency admissions did not vary between threshold Index NEWS2 scores ≥3 to ≥6. Although just one means by which to define an optimal criterion for discrimination, equivalence of the J statistic across a range of scores emphasises the fact that other factors must be accounted for, including outcome prevalence and response cost, even in Day 1 emergency admissions. The finding of little change in sensitivity for NEWS2 scores in the range 4 to 6, is reminiscent of the findings of Pimentel and colleagues comparing NEWS and NEWS2 in the whole in-patient population (17). They showed NEWS2 performed somewhat less well than NEWS in this range, most notably in the majority of patients not at risk of hypercapnic respiratory failure.

Since patients within 24 hours of admission only constitute 21% of bed occupancy at midday, definition of an optimal key trigger threshold is even more important beyond the first day. For example, day 2-14 post-admission, the Index NEWS2 associated event rate was 1.95/1000 patients/day; the false negative rate for a threshold Index NEWS2 score ≥5 = 1.13/1000 patients/day and for an Index NEWS2 score ≥6 = 1.36/1000 patients/day. This single step increase in alert threshold was associated with the daily number of patients requiring a clinical response falling from 100/1000 to 53/1000 patients present at midday.

Health economic evaluation of EWS implementation is sparse (19), we therefore used Simmes and colleagues 2009 cost of ‘medical emergency team’ assessment in Holland (10). Allowing just one assessment per patient reaching a NEWS2 threshold score in every 24-hour period post-admission, generated a cost of €26,463 for every additional composite outcome event in which a response was triggered by a threshold NEWS2 score ≥5 vs ≥6, for the patient population ≥24 hours post admission. This analysis reduced all NEWS2 readings obtained in a day into a single maximum to define satisfaction of a threshold and then applies test performance derived from a single daily Index NEWS2 score. Although this is a simplification, any other consistent set of assumptions increase the incremental costs. Full health economic evaluation is further complicated by the fact that there are additional opportunities to identify the deteriorating patient, including routine clinical evaluation or later NEWS2 triggering at a higher threshold. Also, the benefits of intervening following current key threshold NEWS2 scores have still to be defined (11, 12, 20). The opportunity cost is perhaps easier to represent, specifically the reassignment of resource from one patient group to another on the basis of a screening test with evident limitations. Applying the assumptions made by Simmes and colleagues (10), the additional human resource required to meet a threshold NEWS2 ≥5 vs ≥6 is 18 whole time equivalent healthcare professionals/1000 beds occupied at QEHB. For the reasons described above, this represents a lower limit of resource. The significant incremental cost of triggering at this step, seem also to apply to Day 1 of emergency admission, although in the context of admission units, this cost might be offset by the presence of pre-existing focussed resource.

These findings are also relevant to recent NHSE recommendations on the detection of sepsis, designating NEWS2 ≥5 as the threshold for prompt clinical assessment by a senior decision maker (2, 21). This recommendation was derived from data unsuited to the development of a screening test threshold. It was based upon the evaluation of a non-standard outcome in a small sub-population of emergency department attendances already diagnosed with infection and with features of sepsis (22). It was further supported by extrapolation of qSOFA (23, 24), which is a sepsis mortality indicator rather than a population screening tool. The data presented here, identify the real-world consequences of such recommendations, specifically the potential to divert significant senior clinical resource, based upon an unvalidated test.

In emergency admissions, the performance of Index NEWS2 on day 1 was lower than anticipated from the literature on NEWS and NEWS2 (3-7). This could result from methodological differences in other studies, such as inclusion of all available NEWS2 scores, exclusion of day-case admissions and the need to infer end of life pathways. For example, in our study the inclusion of patients with a DNACPR order for Day 1 emergency admissions, resulted in a c-statistic = 0.87. Also, post-hoc analysis of Day 1-14 emergency admissions that included all NEWS2 scores, generated a c statistic = 0.86 compared to 0.80 using the Index NEWS2; data which imply an association between NEWS2 performance and the frequency with which it is measured. A finding that justifies the design of our primary analysis, in which a single score is used per day. This finding is relevant to interpretation of the existing literature and specifically its application to rules-based clinical response recommendations.

Our single centre study has the advantage that existing SEWS based alerting protocols remained in use, so it is unlikely that these findings are confounded by alerting strategy, through a response-based reduction in event rate. Of the 5089 Index NEWS2 scores = 5, only 375 met the current SEWS alerting threshold ≥4, in whom there were two outcome events.

A single centre design does have limitations with respect to generalisation to other populations, but our findings are supported by data within the report of Kovacs and colleagues in another, well-studied, large acute hospital. They report comparable or higher activity arising from a NEWS ≥5, with similar changes in clinical response requirements with each step change in NEWS threshold (7). In 2012, Greengross and Beaumont described similar unpublished data from multiple users of the VitalPAC system, concluding that ‘The RCP recommendations (on NEWS) may not be sustainable (across an entire in-patient population)’ (25). Our data suggest that this is not simply an issue of capacity but potential jeopardy in the redirection of resource. In an acute admissions unit in which event rates are high and resource already focussed, value may be efficiently realised from a well calibrated risk assessment that allows healthcare professionals to ‘track’ patients. This is a significantly different task to the efficient discrimination required of a screening test to ‘trigger’ a clinical response in the whole in-patient population.

In conclusion, there is a need for further development of EWS’s and recommended clinical response thresholds that can be implemented at reasonable resource cost across the in-patient population. On the basis of these data, it is unclear whether the implementation of a key threshold NEWS2 score ≥5 across the entire in-patient population is feasible or warranted within any given resource. Single step changes in recommended clinical response thresholds have significant implications for resource allocation and their definition require careful consideration. Given existing reports of NEWS and NEWS2 performance, our findings are highly likely to be relevant to other acute hospitals.

## Data Availability

The anonymised participant data and a data dictionary defining each field will be available to others through application to PIONEER, the HDR-UK Health data Hub via the corresponding author. The data will be available upon request and following approval of a process to ensure ethical data governance and through a data access agreement. Please contact the corresponding author for details.

## Copyright and License for publication

The Corresponding Author has the right to grant on behalf of all authors and does grant on behalf of all authors, a worldwide licence to the Publishers and its licensees in perpetuity, in all forms, formats and media (whether known now or created in the future), to i) publish, reproduce, distribute, display and store the Contribution, ii) translate the Contribution into other languages, create adaptations, reprints, include within collections and create summaries, extracts and/or, abstracts of the Contribution, iii) create any other derivative work(s) based on the Contribution, iv) to exploit all subsidiary rights in the Contribution, v) the inclusion of electronic links from the Contribution to third party material where-ever it may be located; and, vi) licence any third party to do any or all of the above.

## Patient and Public involvement

302 patients and public members were consulted as to the use of health data to improve the care for people with acute, unplanned illness. The theme of acuity scores were discussed in a working group and agreed to be a priority for patients. The results of this paper will be disseminated through the PIONEER patient and public group.

## Contributing statement

Pankhurst, Gkoutos and Ball designed the study. Gyves, Evison, Gallier curated the health data and conducted the analysis. Sapey and Ball wrote the paper. Pankhurst, Gallier, Gkoutos contributed to manuscript revision. All authors approved the final version.

## Transparency declaration

E Sapey (the manuscript’s guarantor) affirms that the manuscript is an honest, accurate, and transparent account of the study being reported; that no important aspects of the study have been omitted; and that any discrepancies from the study as planned (and, if relevant, registered) have been explained.

This study was approved by the UHB audit and quality improvement system (CARMS-15850)

The study was funded by Health Data Research -UK as part of PIONEER but the funder did not direct or contribute to any research outputs and researchers developed, conducted and wrote up this report independent to the funder

The study was sponsored by University Hospital Birmingham NHS Foundation Trust

